# The return of human rabies: A foretold case in Latin America Through the Lens of One Health

**DOI:** 10.64898/2025.12.31.25343275

**Authors:** Ricardo Castillo-Neyra, Lizzie Ortiz-Cam, Elvis W. Díaz, Jorge L. Cañari-Casaño, Sergio E. Recuenco, Valerie A. Paz-Soldán

**Author notes:** Corresponding Author (RCN) 423 Guardian Drive Room 715, Blockley Hall Philadelphia, PA 19104-6021. These two authors contributed equally to this work (Ricardo Castillo-Neyra, Lizzie Ortiz-Cam).

## Abstract

**Background:** Despite ongoing rabies control efforts in Arequipa, Peru—including mass dog vaccination campaigns and reactive ring interventions—the region has failed to reduce the number of rabid dogs, leading to the first reported human dog-mediated rabies case after 8 years. The gaps in the rabies control program and the complex dynamics among stakeholders are unknown.

**Objective:** To integrate epidemiologic, socio-ecological, and policy data to identify the factors contributing to the 2023 human rabies case and propose strategies to make this a ‘never event’.

**Methods:** We used stakeholder mapping and field quantitative and qualitative data to identify the roles and connections of key actors in rabies control, identifying gaps in their functions. We then applied the Swiss Cheese model to characterize the defense layers against dog-mediated rabies, highlighting critical vulnerabilities across these protective barriers.

**Conclusions:** We identified multiple breaches in the defense against dog-mediated human rabies. Weak surveillance, insufficient dog vaccination, and inadequate management of free-roaming and feral dogs, coupled with bureaucratic inefficiencies, were key gaps. Outbreak responses were delayed and insufficient, and access to post-exposure prophylaxis (PEP) remained limited. Communication breakdowns exacerbated the problem. Systemic issues, such as outdated public health policies, insufficient training of health professionals, and fragmented efforts, further hindered timely exposure response. Dog ecology and demographic factors also contributed to dog rabies spread. These failures in policy, response, capacity, and external factors led to the 2023 human rabies case. Despite awareness of these challenges, the contributing conditions remain unchanged. Eliminating dog-mediated human rabies by 2030 will require targeted interventions, including enhanced surveillance, context-specific policy reforms, stronger community and institutional collaboration, and better management of free-roaming dogs.

## INTRODUCTION

Dog-mediated human rabies remains a major public health threat, causing tens of thousands of deaths annually around the world (Hampson et al., 2015) despite advances in infectious disease prevention and control (Guzman et al., 2022; Schneider & Santos-Burgoa, 1994; Smith, 1996). Latin America has made great progress towards eliminating dog-mediated human rabies under the “Zero by 30” strategy (WHO et al., 2018). However, enzootic foci remain in a few countries including Peru, where repeated reintroductions have complicated control efforts (Castillo-Neyra, Brown, et al., 2017; Recuenco Cabrera, 2019).

In Peru, great advances were achieved by the 2000’s towards the elimination of dog-mediated human rabies through nationwide mass dog vaccination campaigns (MINSA, 2012; Navarro V, 2020). By 2006, 88% of the country was declared rabies-free. However, between 2009 and 2015, dog rabies reemerged in several regions, including the department of Arequipa (Castillo-Neyra, Brown, et al., 2017; Recuenco, 2019). Since the reintroduction detected in 2015, 395 dog rabies cases have been detected with persistent circulation concentrated in urban and periurban areas (Raynor et al., 2021). In 2023, Arequipa reported its first human rabies death in almost one decade. This event occurred despite ongoing mass dog vaccination efforts and reactive outbreak control activities, highlighting critical vulnerabilities in the public health system.

The systemic nature of these failures remains poorly understood. Existing literature identifies risk factors such as low vaccination coverage, limited PEP availability, weak intersectoral coordination (Benavides et al., 2019; da Silva et al., 2022; Gianella et al., 2020; Navarro V et al., 2007; Roberti et al., 2024), and dog ecology shaped by socioeconomic factors (Castillo-Neyra, Zegarra, et al., 2017; De la Puente-León et al., 2020; Moran et al., 2022; Raynor et al., 2020; Subedi et al., 2022). However, few studies have examined how these factors interact within a real-world event to produce a fatal outcome.

In this study, we analyze the 2023 human rabies case in Arequipa using an integrated One Health approach (Cleaveland et al., 2014). We combine quantitative and qualitative field data with stakeholder mapping and apply the Swiss Cheese model (Reason, 1990; Stein & Heiss, 2015) to identify the specific system breaches that permitted transmission Our objective is to understand how these failures aligned, to illuminate systemic gaps, and propose strategies to prevent similar events in the future.

### Research in context

#### Evidence before this study

Dog-mediated human rabies remains a significant public health threat globally. In Latin America, substantial progress has been made toward the elimination of this disease in alignment with the ’Zero by 30’ strategy (zero human dog-mediated rabies deaths by 2030). While the region reported hundreds of human cases annually in the early 1980s, incidence has since declined by approximately 98%. Countries or subnational areas that continue to report human rabies are often perceived as having weak health systems. However, this systemic weakness has yet to be comprehensively analyzed and characterized to identify the persistent gaps, needs, and challenges necessary to make human rabies a ’never event’ in the Americas.

We searched PubMed, Scopus, SciELO, Google Scholar and national public health reports for studies published between January 2000 and January 2025 using the terms “dog rabies Latin America,” “human rabies Peru,” “rabies control programs,” “dog-mediated rabies,” “massive dog vaccination” and “One Health rabies.” We included studies in English and Spanish and focused on peer-reviewed articles, policy reports, and field studies related to rabies reemergence, outbreak responses, vaccination coverage, and One Health approaches in Latin America. Previous research identified key challenges to rabies control in the region, including inconsistent vaccination coverage, weak surveillance, limited access to post-exposure prophylaxis (PEP), and poor coordination between health, veterinary, and local authorities. Despite recognition of these barriers and existing initiatives such as the “Zero by 30” , human rabies cases continue to emerge, particularly in areas with reintroduced canine rabies, such as Arequipa, Peru.

#### Added value of this study

This study provides the first in-depth, multidisciplinary analysis of the 2023 human rabies case in Arequipa, Peru—an urban setting where dog rabies had been previously reintroduced but never led to a human fatality. By integrating stakeholder mapping, community data, policy review, and by applying the Swiss Cheese model, we identify the specific operational, structural, ecological, and policy-level failures that contributed to this preventable death. Our study uniquely illustrates how systemic gaps, including insufficient dog vaccination coverage, delayed outbreak response, inadequate management of free-roaming dogs, and weak intersectoral coordination, aligned to create conditions for the virus to reach the human population. This analysis situates the event within a broader socio-ecological and policy context, offering a novel interdisciplinary lens.

#### Implications of all the available evidence

Despite substantial regional progress towards the “Zero by 30” goal, this case illustrates that systemic vulnerabilities remain deeply embedded in local and national public health structures. The 2023 human rabies case should serve as a wake-up call. Integrating One Health principles, improving intersectoral communication, ensuring timely PEP availability, and strengthening surveillance and vaccination campaigns are critical to prevent similar tragedies. Our findings provide a practical framework for countries with persistent dog rabies to conduct failure analyses and redesign multisectoral rabies control strategies to prevent future deaths.

## METHODS

### Study design

We conducted a mixed-methods, One Health investigation to examine the systemic failures that enabled the 2023 dog-mediated human rabies case in Arequipa, Peru. The study integrated epidemiologic, socio-ecological, and policy data. Stakeholder mapping was used to identify institutional roles and interactions, and the Swiss Cheese model served as an analytical framework to characterize vulnerabilities across multiple layers of the rabies prevention and control system.

### Conceptual frameworks

#### Stakeholder mapping

Stakeholder mapping can be used to identify key stakeholders, categorize their level of support and influence for a given initiative, analyze stakeholder relationships and dynamics, and evaluate their potential to serve as allies or mitigate potential risks to successful project implementation (Brugha & Varvasovszky, 2000; Clarkson, 1995; Crosby, 1991). As part of a larger project and before applying the Swiss cheese model, we conducted stakeholder mapping to identify key actors in rabies control and understand their roles within the system and their connections. We categorized stakeholders based on their involvement in rabies control, influence on decision-making, and capacity to contribute to prevention and response activities. This process assessed how these actors interacted within the rabies control framework and examined broader socio-ecological factors influencing rabies transmission.

#### Swiss Cheese Model Analysis

We applied the Swiss Cheese Model(Reason, 1990; Stein & Heiss, 2015) as a framework for understanding how multiple layers of defense, which are represented as cheese slides, failed to prevent the transmission of rabies. This model is particularly useful for identifying systemic vulnerabilities and areas where protective measures may have been inadequate.

We first identified the key defensive barrier groups in place at the time of the rabies transmission, including policies, outbreak response activities, system capacity building and program/process evaluation. We then determined the specific defensive barriers or *slices* that are involved within these groups and examined for weaknesses or *holes* that allowed the virus to bypass the defense layers. Each barrier was systematically analyzed for its effectiveness, with particular focus on how organizational, logistical, and socio-economic factors may have contributed to these failures. To support our assessment, we conducted a comprehensive review of qualitative and quantitative data collected from the field, including vaccination coverage reports from the rabies victim’s district and surrounding areas, rabies surveillance records, outbreak detection and response times, and documentation of response activities, such as the availability and administration of post-exposure prophylaxis (PEP).

### Data sources

#### Surveillance Data

We reviewed dog rabies surveillance records from 2015-2023, including samples submitted for diagnostic testing through passive and active surveillance. Active surveillance included field investigations, carcass sampling along dry water channels, and response to community notifications of suspect dogs. Human bite records, outbreak reports, and PEP administration data were also obtained from regional health authorities.

#### Dog vaccination coverage

Mass dog vaccination coverage data for Arequipa and the Chiguata *microred* (healthcare operational unit serving an area approximately equivalent to a district) were compiled from Ministry of Health (MOH) reports. Coverage estimates were evaluated relative to recommended targets for achieving herd immunity. Dog population estimates were reviewed to assess the accuracy and consistency of denominators used to calculate coverage.

#### Mixed-Methods community data

##### Door-to-door (D2D) surveys

Since 2016, we have conducted D2D structured surveys in urban and periurban communities of Arequipa, with a pause only in 2020-2021. The annual number of surveyed households has fluctuated from ∼2,000 to ∼8,000. The surveys were conducted immediately after the annual mass dog vaccination campaigns (MDVC) and the survey’s questions include themes such as dog ownership practices, rabies vaccination status, SES, and family’s migration history. All surveyed houses were georeferenced as well as the vaccination sites during the dog vaccination campaign. Description of the study communities and the data collection approaches are described elsewhere (Castillo-Neyra et al., 2019).

##### Focus groups with communities

We conducted 8 focus groups (FG) in pre-pandemic years and 7 focus groups post-pandemic with dog owners and other community members. We developed FG guides to cover five topics: dog ownership, dog ecology, barriers to and facilitators of dog vaccination, and preventive practices following a dog bite (i.e., post-exposure prophylaxis, seeking healthcare). We recruited participants in urban and periurban areas from different socioeconomic status and representing a range of ages, gender, dog ownership, and dog vaccination status (i.e., we aimed to include both dog owners who had and had not participated in the vaccination campaigns). We used a mixture of deductive and inductive codes to analyze the data. Sampling techniques, recruitment approaches, and data collection and analysis details are described elsewhere (Castillo-Neyra, Brown, et al., 2017; Castillo-Neyra et al., 2020, 2025).

##### Focus groups with public health authorities

We conducted a focus group with key official stakeholders including representatives from the surveillance program, regional laboratory coordinators for rabies diagnosis, health promotion, health communication, immunizations (including cold chain), implementation of the mass dog rabies campaigns, and regulatory authorities of the dog rabies elimination program (n=11). Together, these authorities dictate local rabies control guidelines, request budgets, approve local plans, provide technical support for the control activities, plan the city-wide dog rabies programs, manage and submit data to the national level, and supervise the program’s activities. Sampling, recruitment, data collection and analysis are described elsewhere (Castillo-Neyra et al., 2025).

##### Group Model Building workshop with dog rabies elimination program implementers and authorities

We conducted a day-long GMB workshop with 69 implementers and authorities in charge of surveillance, mass dog vaccination campaign implementers, and community awareness and engagement. They were divided in 15 small working groups to discuss barriers and challenges for dog rabies elimination. The output from these working groups was then discussed by the larger group with our field research team observations. The methodological details of this GMB workshop are described elsewhere (Castillo-Neyra et al., 2025).

#### Dog Ecology

In broad terms and based on their level or restriction and dependence on humans, we can think of 4 subpopulations of dogs in Arequipa: well restricted dogs, owned free-roaming dogs, unowned dogs (“strays”), and feral dogs. We have studied their ecology conducting D2D surveys about owned dogs and their population parameters(Castillo-Neyra et al., 2019; Díaz Espinoza, 2019), tracking the movement of free-roaming dogs with GPS collars(Raynor et al., 2020), and conducting monthly observations in the outskirts of the city where feral dogs are present(De la Puente-León, 2024). In addition, regular visits to solid waste landfills, small urban dumps, dry water channels and other places where free-roaming dogs feed, congregate, and mate, provided contextual data to understand social and ecological connections between dog packs and their landscape.

#### Outbreak response

Following the human rabies case was reported, the Chiguata Health Network and our research team conducted targeted visits near the human rabies case and collected data through epidemiological surveys of local residents and health authorities over five consecutive days. Furthermore, thanks to the close collaboration between our research team and the Chiguata’s health authorities, we implemented active surveillance to search for rabies cases along the waterways of this district. Our team carried out this activity two days a week for two months following the human case. During this time, we mapped the area, assisted in collecting samples from potential cases, and observed areas within the dry water channels used for the disposal of organic and inorganic waste.

#### Study Area

San Bernardo de Chiguata (Chiguata hereon) is a relatively new expanded peri-urban area (Figure 1) with rapid population growth, characterized by agricultural activities, free-roaming dogs, multiple feral dogs inhabiting caves, and proximity to two districts with active dog rabies transmission (CDC-MINSA, 2023b).

**Figure 1.**
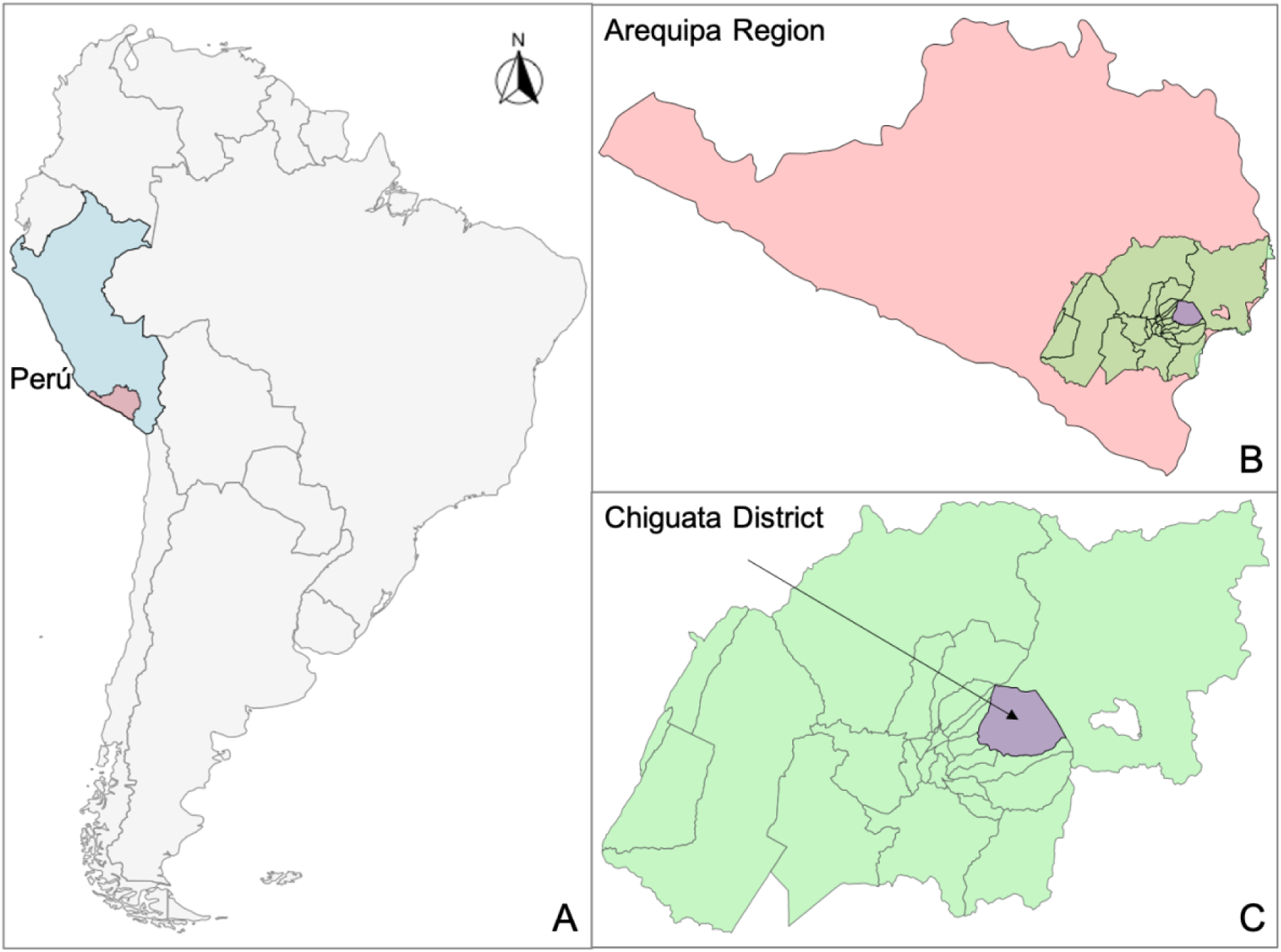
Peru, country which reported the fatal human rabies case in 2023 (A). Arequipa region, a canine rabies endemic area in Peru (B). Chiguata district, located within Arequipa city (C).

#### Ethics

Our research team obtained IRB approval from University of Pennsylvania (UPENN), Universidad Peruana Cayetano Heredia (UPCH) and Tulane University for conducting focus groups and surveys (IRB # 823736 and 850695, 65369 and 207735, 606720 and 2022-1134, respectively), and workshops and observations of dog rabies vaccination campaigns in Arequipa (IRB 823736, 65369, 606720, respectively).

## RESULTS

### Stakeholder mapping

The results of our stakeholder mapping highlight the complexity and interdependence of the various actors involved in rabies control in Peru. Figure 2 presents a detailed overview of the key stakeholders, their roles, and the relationships between them. The MOH plays a central role in establishing regulations, goals, and providing necessary resources such as the main budget, vaccines, and syringes. Other governmental bodies, including the regional health management team and local government, support these efforts by coordinating logistics, outbreak control activities, and surveillance.

**Figure 2.**
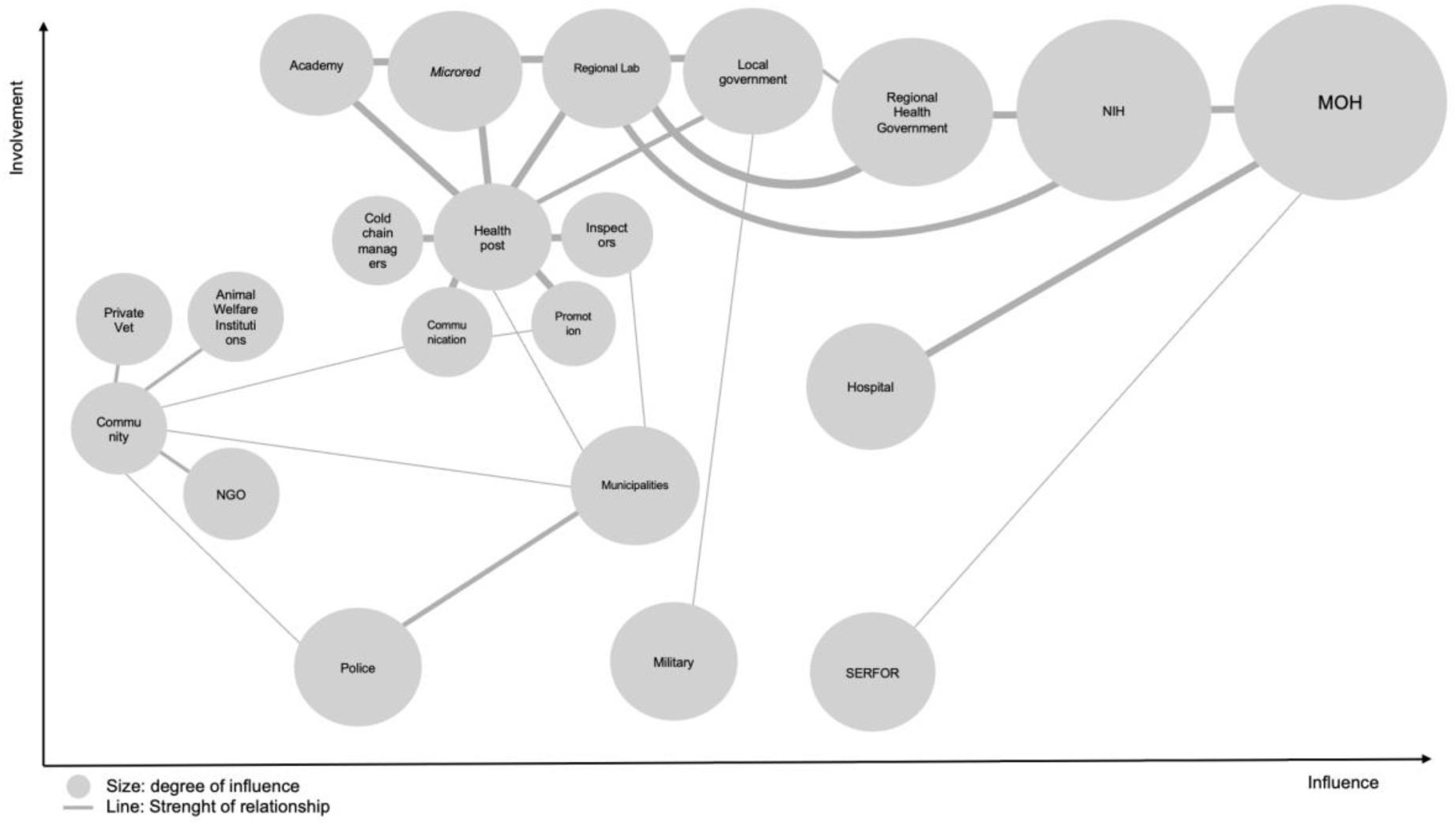
Stakeholder map for dog rabies control in Peru.

Key entities such as municipalities, the military, and the police provide critical logistical and security support during vaccination campaigns and outbreak response activities, ensuring order. Non-governmental organizations (NGOs), private veterinarians, and community members also contribute to rabies control, though their roles are more focused on promoting awareness, animal welfare, and vaccination services.

### Gaps identified by Swiss Cheese Model analysis

Building upon the stakeholder mapping, and using the Swiss Cheese Model analysis, we identified the key defensive barriers (*slices*) against the rabies virus and the gaps or failures (*holes*) that allowed the transmission of the rabies virus (Figure 3). We classified these key defensive barriers into four large groups: the **Policies** barrier group, where *slices* included surveillance, mass dog vaccination campaigns (MDVC), dog population management, and garbage control. The **Outbreak Response Activities** group, which included as *slices* suspect case notification, outbreak control activities, and PEP. In the **System Capacity Building** group, health professional training and community education plans were identified *slices*, while the **Ecological & other factors** group included the *slices* geographical features, and migration patterns. Here, we provide an in-depth analysis of each *slice* of cheese within the Swiss Cheese Model, in the context of the human rabies case.

**Figure 3.**
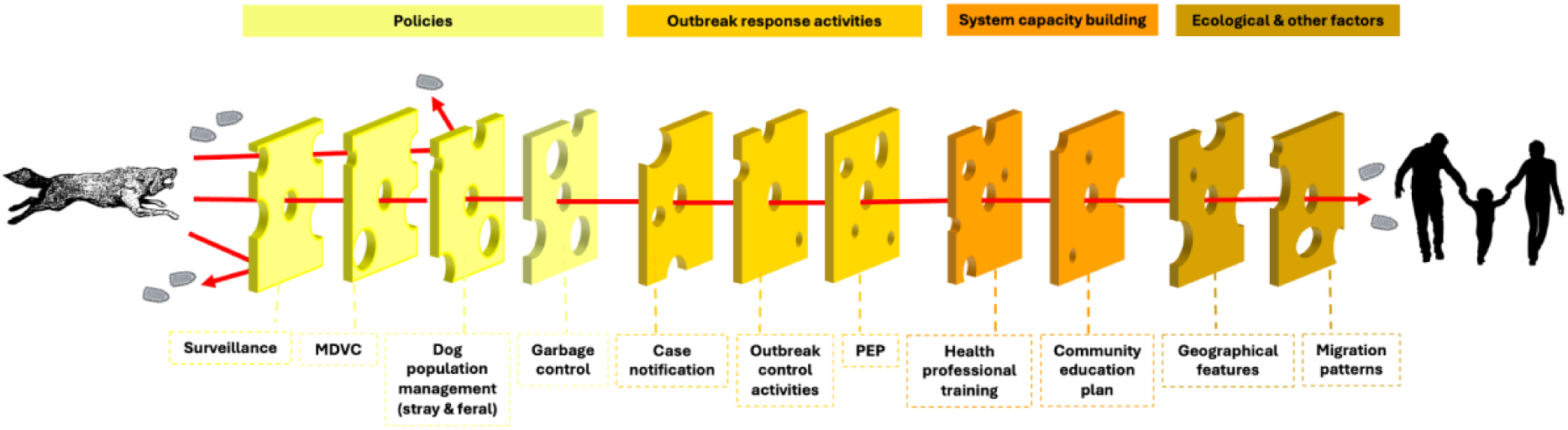
Swiss Cheese model analysis for rabies.

### Policies

#### a. Surveillance

From 2015 to 2020, the number of dog samples collected by the MOH steadily decreased by nearly 80% (from 737 to 153) (Figure 4). Although sample numbers increased again after 2021, partly due to our team’s active surveillance, canine rabies continued to be underdetected. In Chiguata, no structured monitoring of dog bites, free-roaming dogs, or suspect cases was in place. Regarding human cases, no search of human deaths associated with encephalitis was conducted. Field interviews confirmed that bite incidents were neither systematically reported nor linked to public health follow-up.

**Figure 4.**
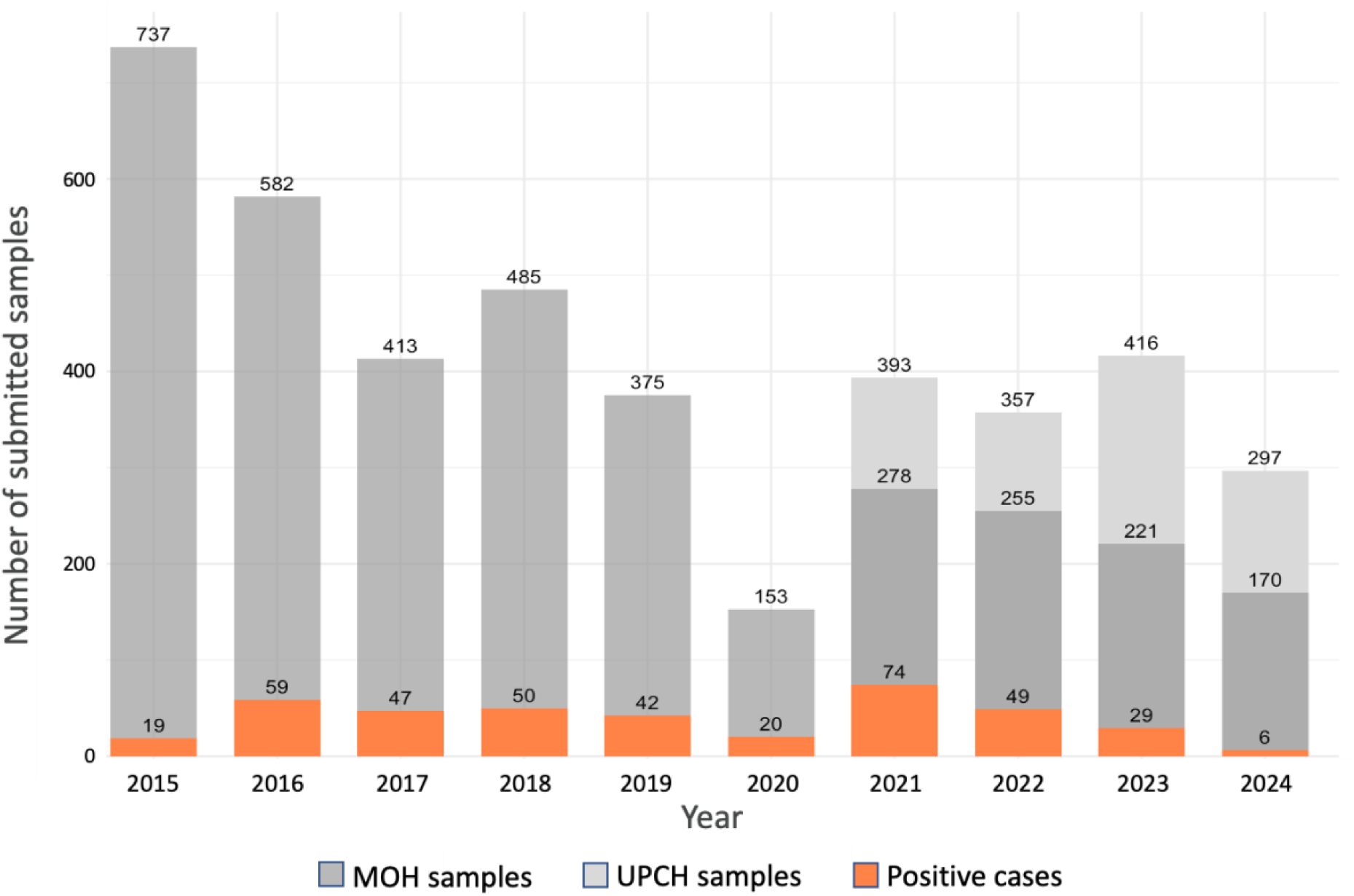
Dog rabies cases identified and submitted samples in Arequipa City, 2015 to August 2024.

#### b. Mass dog vaccination coverage

In Arequipa, we have observed and supported these campaigns since 2015. Initially, campaigns were conducted over one or two weekend days, but more recently, they have been staggered over several months(Bellotti et al., 2025). Vaccination is offered free of charge in outdoor settings from 8 a.m. to 1 p.m., and participation is voluntary. The MOH uses a cell-culture-based vaccine and is focused solely on dogs with owners (MINSA, 2017). Campaign communication and promotion are conducted at the local level. Vaccination point locations are determined a few days in advance; some are relatively consistent year to year (e.g., the entrance of a health post), while others may be selected the day of the campaign. Teams may relocate during the day from areas of lower demand to higher demand. The dog population—used as the denominator for estimating coverage—is calculated using the human-to-dog ratio method. This ratio varies significantly by geography(Downes et al., 2013), and in Arequipa, it has evolved from 10:1 in 2015 to 6:1 and, more recently, to more refined estimates(Castillo-Neyra et al., 2019). As a result, accurately assessing vaccination coverage remains a moving target.

Figure 5 reveals fluctuation in the vaccination coverage in Arequipa since 2015, showing that the minimum recommended coverage to achieve herd immunity is not reached in the majority of the years. This is due to quantitative and qualitative factors, which have been previously studied and published (Castillo-Neyra, Brown, et al., 2017; Castillo-Neyra et al., 2019).

**Figure 5.**
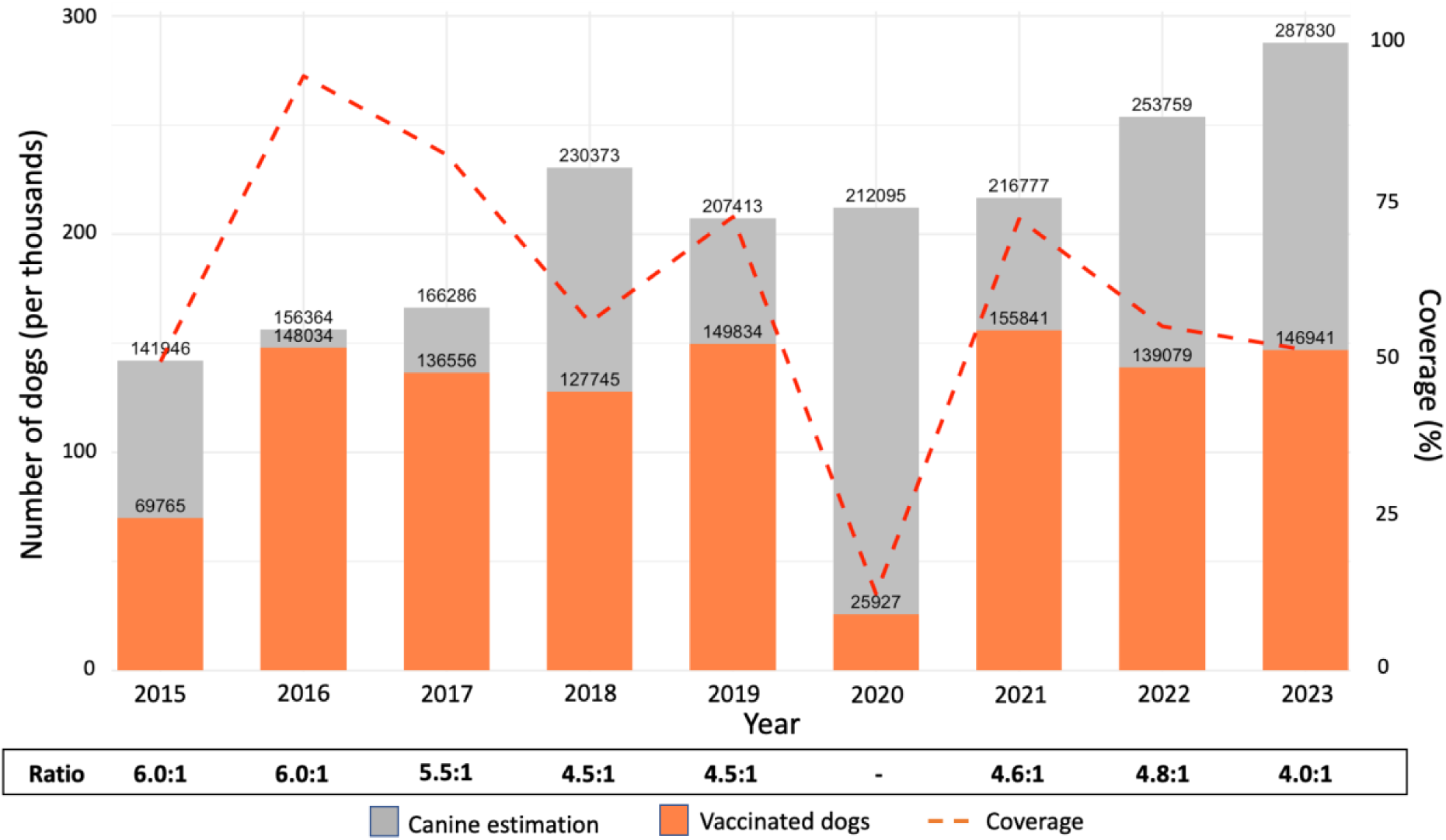
Dog vaccination coverage in Arequipa, Perú 2015 - 2023.

The Chiguata *microred* carries out its rabies vaccination campaigns on an annual basis, primarily targeting accessible dogs, defined as owned dogs that can be restrained by their owners. In 2021, it reported more than 80% vaccination coverage; in 2023, it carried out its vaccination campaign in September and vaccinated 68.5% animals up to that month (Table 1).

**Table 1.** Estimates used by Chiguata *microred* for local public health activities (2021-2023) Source: Arequipa - Caylloma Health Red. September 2023 \**More than 100% coverage was reported, which indicates an underestimation of the canine population*.

| Year | Human population | Ratio H:D | Dog population | Vaccinated dogs | MDVC coverage |
| --- | --- | --- | --- | --- | --- |
| 2021 | 6208 | 3.4 | 1800 | 1500 | 83.3% |
| 2022 | 3266 | 2.5 | 1306 | 1478 | 113.2%* |
| 2023 | 4629 | 2.5 | 1852 | 1269 | 68.5% |

#### c. Dog population management

Municipalities are responsible for dog population control, yet field assessments revealed: i) the presence of free-roaming dogs that routinely moved between households (Figure 6B), streets, and water channels; ii) aggressive behaviour from free-roaming dogs towards residents, their pets and foreign visitors; iii) and unregulated feeding of unowned dogs by residents, reinforcing established roaming patterns (see item j).

**Figure 6.**
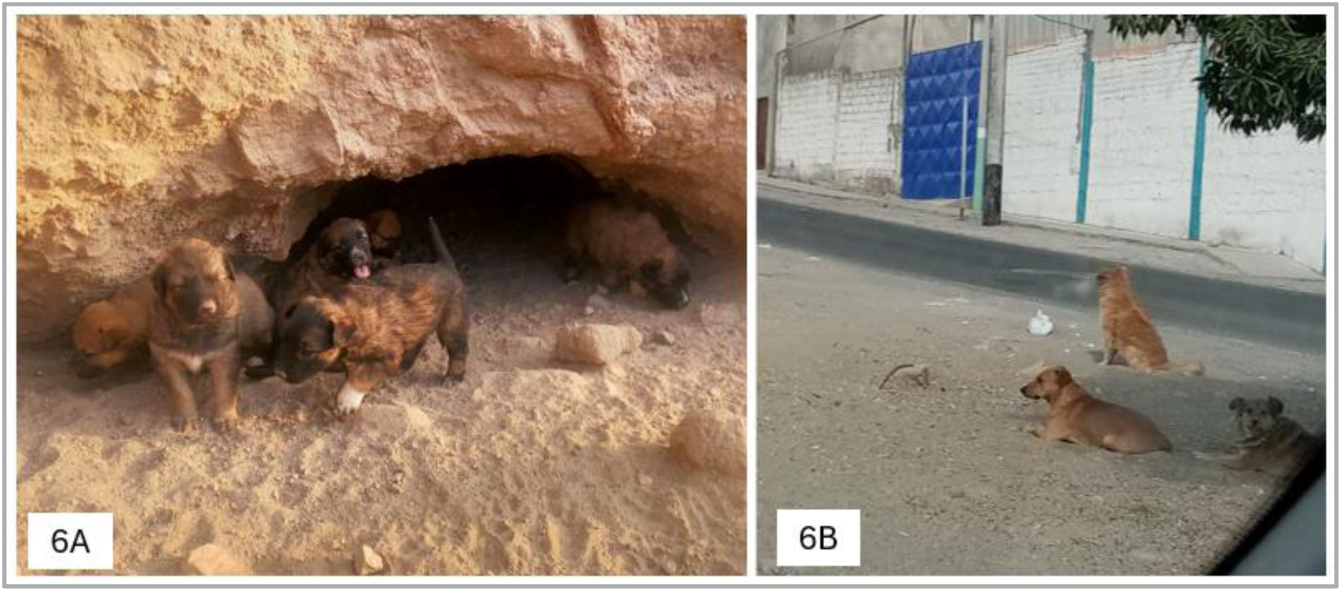
Dog population in Arequipa, Peru. (A) Caves used by feral dogs, active. (B) Presence of free-roaming dogs in the city.

But in addition to the free-roaming urban dog population, there is a potentially bigger problem: feral dogs (Piperis & Huisa-Balcon, 2024). Within the study area, our field team observed five cave areas, and an area with a litter of 6 puppies (Figure 6A) whose location was at a five-minute walking distance to the victim’s house. Moreover, we were able to identify other characteristics of the site which we have represented in Figure 7. A total of 15 free-roaming dogs, 12 caves, three areas of informal waste disposal sites and four agricultural areas.

**Figure 7.**
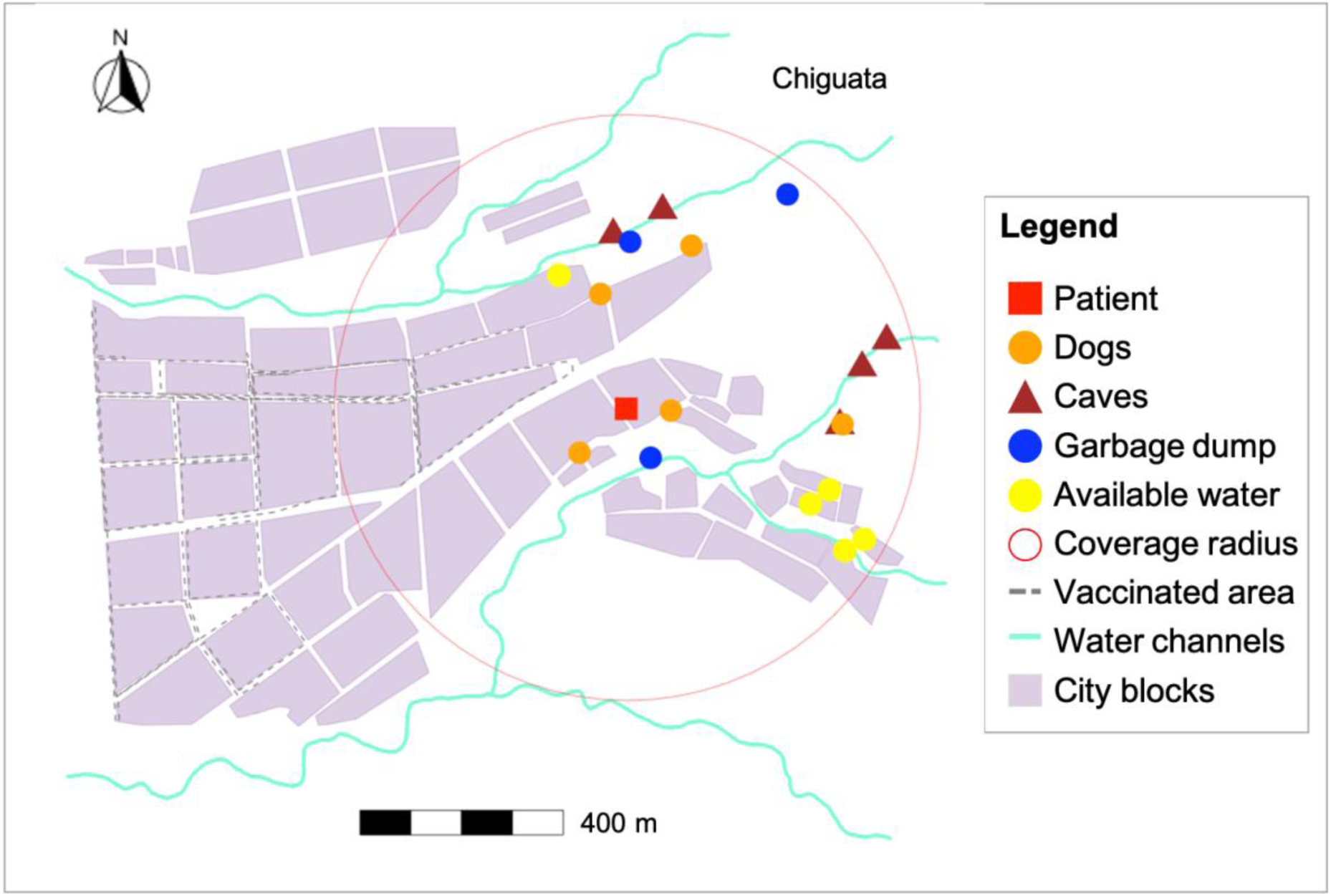
Characteristics of the study area.

#### d. Waste management

In periurban Chiguata, substantial organic and inorganic waste accumulated along dry water channels (*Torrenteras*) and in nearby areas (Figure 8). Household waste disposal was irregular, and many residents deposited agricultural and domestic waste directly into the channels. These waste sites provided a persistent food source, enabling free-roaming and feral dogs to congregate, reproduce, and travel along the channels.

**Figure 8.**
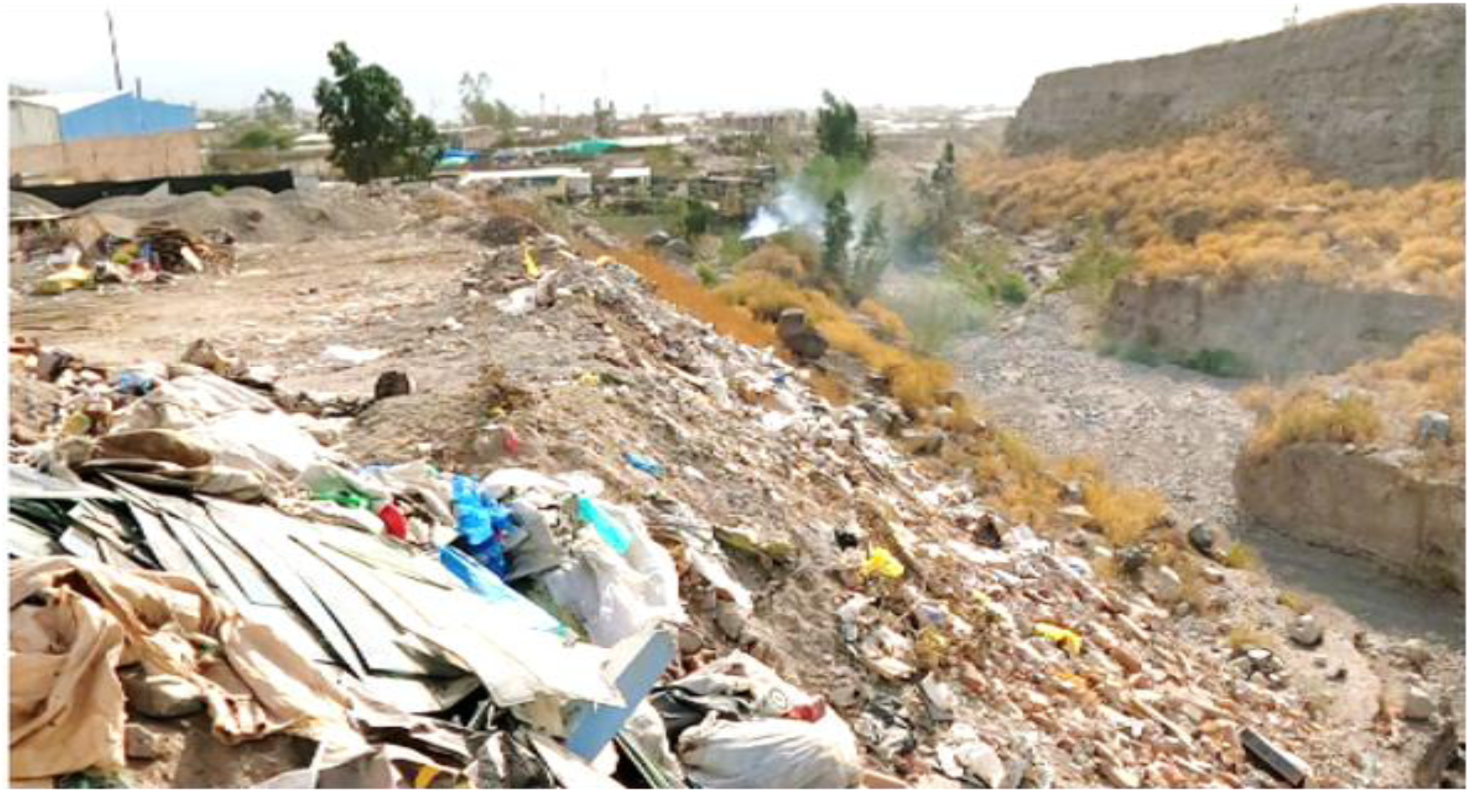
*Torrentera* during the dry season, with waste disposal in it.

### Outbreak response activities

#### e. Case notification

Effective case notification is a critical layer in the Swiss cheese model of outbreak response, as timely reporting of suspected cases enables swift action to prevent further transmission. However, our analysis revealed significant gaps in this layer. Approximately 62% of rabies cases were reported late, which delays public health interventions such as ring vaccination, contact tracing, PEP administration, and quarantine measures for exposed animals.

Additionally, some rabid dogs exhibiting unmistakable clinical signs of rabies were never reported. Instead, they were clandestinely buried by their owners, likely due to fear of stigma, misinformation, or a lack of awareness about the importance of notifying authorities. This practice not only hampers the official case count but also undermines efforts to identify potential exposures and assess the extent of an outbreak.

#### f. Outbreak control activities

Standard outbreak control guidelines in Peru require a 500m radius response over two months. However, responses typically lasted 1-3 days, with smaller operational areas due to personnel shortages and other competing priorities.

For the 2023 human case, health authorities performed an intensive 5-day emergency response. Two cold-chain containers were prepared, containing 60 and 65 doses of rabies vaccine, respectively. The team formed two groups to conduct ring vaccination in high-risk areas identified during the initial investigation. In addition, five individuals from the Chiguata *microred* were assigned to conduct epidemiological surveys, which included finding and documenting recent cases of dog bites, identifying potential sources of exposure, meaning identifying the suspect rabid dog, and ensuring that at-risk individuals were receiving appropriate medical attention.

#### g. Rabies post-exposure prophylaxis (PEP)

PEP is a life-saving treatment administered after potential exposure to the rabies virus, typically through an animal bite (CDC, 2024). According to the Peruvian Technical Health Standard for the Prevention and Control of Human Rabies (OPS & MINSA, 2015), all individuals who have been bitten or had contact with suspected animals and lived in areas where dog rabies is enzootic, prophylaxis should be initiated immediately upon seeking medical care. Despite these guidelines, ensuring consistent access to PEP can be challenging in some health centers. In our case analysis, the Chiguata health post had no rabies vaccine at the time of the incident; the victim was referred to a different health facility due to jurisdictional divisions, 11km away (Figure 9), creating delays and confusion.

**Figure 9.**
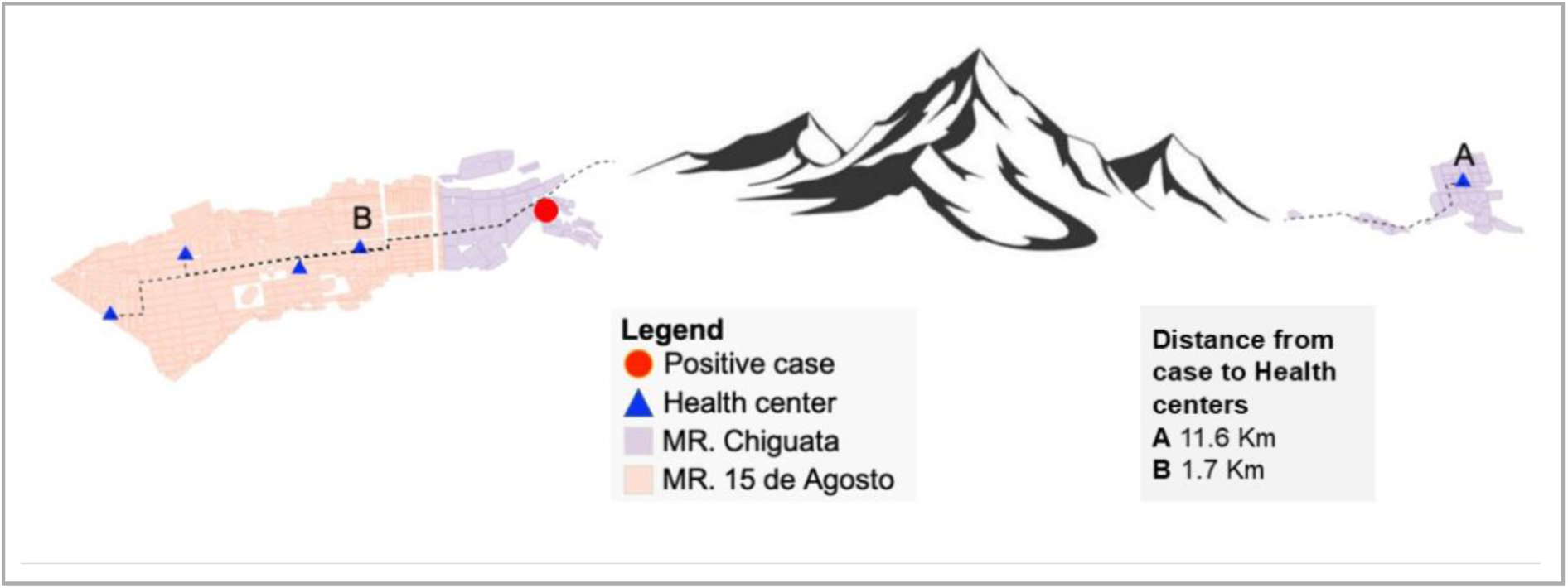
Distribution of health centers in *microreds*, and distance from the case residence to those centers.

Additionally, interviews indicated that PEP shortages occurred in other health centers following a surge in bite reports after media coverage. Vaccines were administered to all patients, regardless of the level of exposure or risk.

### System capacity building

#### h. Health professional training

During the victim’s multiple visits to health centers (Figure 10), rabies was not initially included in the differential diagnosis. Symptoms were attributed to psychiatric or anxiety disorders (CDC-MINSA, 2023a; Gerencia Regional de Salud de Arequipa, 2023; Gobierno Regional de Salud de Arequipa, 2023), delaying appropriate treatment and investigation. Our analysis identified two underlying factors: i) limited training in rabies clinical recognition, and ii) lack of routine inquiry about animal exposure, especially in periurban communities where dog bites are normalized. Only when neurologic symptoms worsened did providers suspect rabies, by which time clinical progression was irreversible.

**Figure 10.**
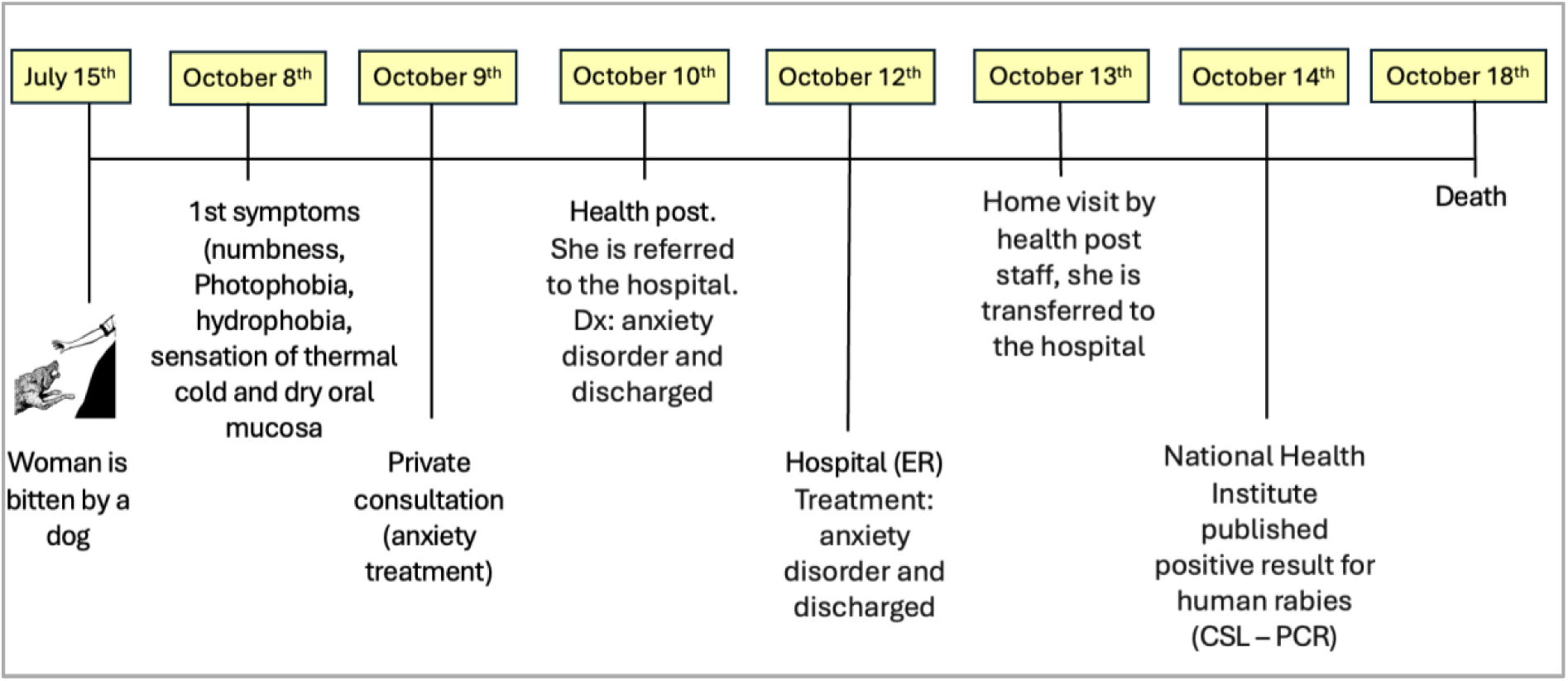
Human rabies case Timeline, Arequipa - 2023.

#### i. Community education plan

Prior to the incident, Chiguata lacked structured rabies education initiatives, which contributed to low unawareness of the need for immediate wound care, the importance of seeking urgent medical attention after a dog bite, and the risk associated with free-roaming dogs. The victim did not report the dog bite, a pattern commonly described in our focus groups, reflecting widespread underestimation of rabies risk. After the case, community-wide awareness campaigns were initiated through door-to-door visits.

### Ecological and other factors

#### j. Geographical features

Dry water channels are a defining ecological feature of Arequipa, which serve as efficient corridors for dogs to move between urban and peri-urban areas (Raynor et al., 2020). Such pathways complicate rabies control, as infected animals can use them to travel undetected, increasing the risk of rapid virus transmission across different dog populations. In addition to facilitating movement, field assessments showed that the channels offer shelter and access to food waste, attracting feral dogs that are particularly challenging to monitor and vaccinate.

#### k. Human migration

Previous studies demonstrate seasonal and economic migration between Arequipa and Puno, where rabies remains endemic. Dog movement driven by human migration along these routes likely contributed to the 2013 reintroduction (Salazar et al., 2024) and may continue to support sporadic viral incursions. In Chiguata, household mobility and recent in-migration were common. Our previous research has shown that recent migrants may have different patterns of dog ownership, vaccination practices, and health-seeking behaviours compared to long-term residents (Castillo-Neyra et al., 2020; De la Puente-León et al., 2020).

## DISCUSSION

The 2023 human rabies case in Arequipa represents a profoundly preventable death in a country with long-standing experience in rabies control. This case emerged in a district considered protected, despite over a decade of known canine rabies circulation, ongoing dog vaccination campaigns, and PEP availability in urban areas. The absence of recent reported dog cases created a false sense of security that weakened surveillance efforts and reduced vigilance in both humans and animals. Applying the Swiss Cheese model reveals that this failure was the result of sequential gaps across all rabies prevention layers, exposing structural weaknesses that demand a One Health approach (Cleaveland et al., 2014). Effective intervention requires sustained, simultaneous activity in endemic areas and in adjacent ecologically connected areas where infected dogs can move along natural and human-made routes. Such vigilance depends on consistent funding and political commitment (S.a et al., 2018), which are often limited in Peru and other LMICs. Rabies program competes with higher-profile health priorities(Nadal et al., 2022; Raynor et al., 2021), and financial support typically declines when no human cases are reported (Del Rio Vilas et al., 2017; Millien et al., 2015; Recuenco, 2019), regardless of ongoing risks.

MDVCs and PEP are essential yet insufficient when applied without adaptation to local ecological and social contexts (Beyene et al., 2020). The situation in Chiguata district illustrates these limitations. Although official reports indicated high vaccination coverage, neighboring districts had lower coverage, and MDVCs targeted exclusively owned dogs, leaving stray and feral dogs as unvaccinated reservoirs. Peru’s centralized rabies program operates on the assumption that unowned free-roaming dogs are negligible, a premise that does not reflect reality in Arequipa. A large population of unowned dogs inhabits the city’s dry water channels, which serves as ecological corridors linking districts with uneven surveillance and vaccination levels. These dry water channels also provide shelter and access to food waste —in tropical areas, it is estimated that 100 people generate enough refuse to sustain seven stray dogs(Coppinger & Coppinger, 2017)— thereby supporting growing roaming dog populations. In this context, standard MDVCs strategies cannot achieve adequate herd immunity, making tailored vaccination strategies essential for meaningful, sustained coverage.

Free-roaming dog density has been identified as a key predictor of rabies transmission in some settings (Beron et al., 2024), while in others it appears to have limited direct influence on transmission dynamics(Morters et al., 2013). Regardless, larger dog populations impose greater demands on population management efforts for multiple public health purposes. In Arequipa, the absence of systematic dog population management programs, coupled with abundant food waste, creates conditions that strongly favor the persistence and growth of free-roaming dog populations. Addressing these ecological challenges requires tailored interventions, including specialized vaccination strategies and coordinated action efforts involving municipal authorities, agricultural agencies, and private veterinarians (Swedberg et al., 2022).

Surveillance represents another critical weakness. Despite national mandates for canine rabies surveillance, implementation is inconsistent. Arequipa routinely submitted few samples, and no retrospective evaluations were conducted following the human case. Weak surveillance obscures early detection of epizootic expansion and delays timely responses. While modern diagnostics tools can accelerate laboratory confirmation, their utility depends on adequate field sampling, trained personnel, community engagement, and regular sample submission.

Human rabies surveillance faces similar limitations. The Arequipa case mirrors other international examples, such as a fatal case in Tunisia, where clinicians did not investigate animal exposure or consider rabies in the differential diagnosis(Dhaoui et al., 2025). In Peru, insufficient training in syndromic surveillance, poor awareness of rabies risk, and inconsistent access to PEP contribute to delays in care. Systemic monitoring of dog bites, a simple yet powerful early warning tool, is rarely implemented and is often reconstructed only after a human case occurs. Strengthening bite registries, integrating hospital and primary-care data, and training health workers to interpret local epidemiological trends would improve early detection and guide more efficient PEP allocation (Benavides et al., 2019).

Community-based surveillance models from other low-resource settings demonstrate that involving communities in reporting suspect animals and dog bites improves early detection and builds trust with health authorities (Beyene et al., 2020). Integrating local knowledge with national systems could foster a more proactive and sustainable rabies surveillance network. However, effective coordination among health, agriculture, and municipal authorities is a significant challenge, hindered by independent hierarchies and competing priorities.

Several socioeconomic factors further complicate rabies control efforts (Castillo-Neyra et al., 2019). Remote and low-income communities face disproportionate barriers, including distance to health services and weak infrastructure, which delay access to vaccine and PEP (Benavides et al., 2019; da Silva et al., 2022; Gianella et al., 2020; Navarro V et al., 2007; Roberti et al., 2024). Dog ecology is also shaped by factors such as unregulated waste disposal, informal dog ownership, and variable community attitudes toward roaming dogs, creating heterogeneous risk landscapes across Arequipa’s urban and periurban areas (Castillo-Neyra, Zegarra, et al., 2017; De la Puente-León et al., 2020; Moran et al., 2022; Raynor et al., 2020; Subedi et al., 2022).

### A Call to Action: Applying Lessons

The 2023 fatality underscores the urgency of addressing the persistent weaknesses in rabies control strategies. Many required interventions are not costly but depend on clear policies, consistent leadership, and committed action from local and national authorities. However, the structural conditions that enabled this preventable death remain largely unchanged. Strengthening PEP delivery is one actionable avenue. Transitioning to intradermal (ID) vaccine administration could reduce healthcare costs without compromising effectiveness (Lechenne et al., 2017; Swedberg et al., 2022). Though, successful implementation requires rigorous healthcare worker training and robust monitoring. Ongoing research and sustained financing are equally critical. The presence of a research team during this outbreak provides critical real-time insights, highlighting the need to maintain research capacity in endemic regions to refine interventions and inform policy. Funding models must also be reformed; in Mexico, for example, continued support for MDVCs has been contingent on the absence of human rabies cases (González-Roldán et al., 2021), creating a cycle where success threatens program continuity. Breaking this pattern is crucial for maintaining long-term control, thereby preventing future fatalities.

## FUNDING

This project was supported by NIH-NIAID (K01AI139284 and R01AI168291 to RCN) and the Fogarty International Center (D43TW012741 to RCN, LOC, EWD and VPZ). The funders had no role in study, data collection and analysis, decision to publish, or preparation of the manuscript.

## Data Availability

All data produced in the present study are available upon reasonable request to the authors

## Notes

### Competing Interest Statement

The authors have declared no competing interest.

### Author Declarations

Our research team obtained IRB approval from University of Pennsylvania, Universidad Peruana Cayetano Heredia and Tulane University for conducting focus groups and surveys (IRB # 823736 and 850695, 65369 and 207735, 606720 and 2022-1134, respectively), and workshops and observations of dog rabies vaccination campaigns in Arequipa (IRB 823736, 65369, 606720, respectively).

